# Monitoring socioeconomic inequalities across HIV knowledge, attitudes, behaviours and prevention in 18 sub-Saharan African countries

**DOI:** 10.1101/2021.08.24.21262532

**Authors:** Mohamed Hamidouche, Pearl Anne Ante-Testard, Rachel Baggaley, Laura Temime, Kévin Jean

## Abstract

**Objectives:** Socioeconomic inequalities in HIV prevention services coverage constitute important barriers to global prevention targets, especially in sub-Saharan Africa (SSA). We aimed at monitoring these inequalities from population-based survey data in 18 SSA countries between 2010 and 2018.

**Methods:** We defined eight HIV indicators aimed at capturing uptake of HIV prevention services among adult participants. Country-specific wealth-related inequalities were measured using the Relative and Slope Index of Inequalities (RII and SII, respectively) and then pooled using random-effects meta-analyses. We compared inequalities between African regions using the Wilcoxon rank-sum test.

**Results:** The sample consisted of 358,591 participants (66% women). Despite variability between countries and indicators, the meta-analysis revealed significant levels of relative and absolute inequalities in 6 out of 8 indicators: HIV-related knowledge, positive attitudes toward people living with HIV (PLHIV), condom use at last sexual intercourse, participation to prevention of mother-to-child transmission programs, medical male circumcision and recent HIV testing. The largest inequalities were reported in condom use, with condom use reported 5 times more among the richest versus the poorest (RII=5.02, 95% Confidence interval, CI: 2.79-9.05) and in positive attitudes toward PLHIV, with a 32-percentage point difference between the richest and poorest (SII=0.32, 95% CI: 0.26-0.39). Conversely, no significant inequalities were observed in multi-partnership and HIV seropositivity among youth. Overall, inequalities tended to be larger in West and Central vs. East and Southern African countries.

**Conclusions:** Despite efforts to scale-up HIV-prevention programs, socioeconomic inequalities remain substantial over the continuum of HIV primary and secondary prevention in several SSA countries.

## Introduction

Despite a sustained decrease over the last decade, sub-Saharan Africa (SSA) continues to bear a disproportionate burden of the global HIV epidemic, with about two thirds of the worldwide number of HIV infections [1]. Many SSA countries are not on track to meet global targets of 90% reduction of new HIV infections by 2030 compared to 2010 [2,3]. There is however a large body of literature supporting the fact that these targets could be reached by effectively combining existing prevention interventions and tools, including behavioural prevention, condoms, voluntary male medical circumcision (VMMC), pre-exposure prophylaxis (PrEP), HIV testing, and prevention of mother-to-child transmission (PMTCT), alongside increased antiretroviral coverage [4].

HIV risk, knowledge and attitudes, access to services and HIV outcomes vary largely across gender, age, location or social status. Inequalities, including socio-economic inequalities, have been recognized as a contributor in the failure to meet the UNAIDS 90-90-90 testing and treatment 2020 targets [5], and may also be a factor in the disappointing progress in effective prevention coverage. Indeed, socio-economic inequalities have been reported across HIV knowledge [6], HIV testing [7–9], PMTCT [10], and VMMC [11]. However, there has not been to date a comprehensive large-scale assessment of socioeconomic inequalities across the major prevention pillars to guide the focus of future programming to overcome current gaps. Such an assessment would be valuable, though. Firstly, it would allow to identify, within the main HIV prevention pillars, those that could potentially be at specific risk of generating socioeconomic inequalities in HIV infections. Secondly, a monitoring of national HIV prevention indicators focusing on inequalities would potentially allow to identify and then learn from settings that are specifically performant in terms of equity.

In this paper, we used nationally representative surveys conducted across 18 sub-Saharan African countries between 2010 and 2018 to quantify wealth-related inequalities in several indicators aiming at capturing HIV prevention elements.

## Methods

### Study population and data

We analysed data from the most recent Demographic and Health Surveys (DHS) in 18 Sub-Saharan African countries (based on data availability as of November 2020). The country sample was a convenience sample based, and slightly extended, from an earlier research project [9]. The sample included Burkina Faso, Cameroon, Côte d’Ivoire, the Democratic Republic of Congo (DRC), Ethiopia, Guinea, Kenya, Lesotho, Liberia, Malawi, Mali, Niger, Rwanda, Senegal, Sierra Leone, Tanzania, Zambia, and Zimbabwe.

DHS are nationally representative surveys based on a multistage design with households as sampling units. They collect a wide range of demographic and reproduction health indicators among consenting adults that are interviewed face-to-face by trained interviewers using standardized questionnaires [12]. The collected data notably include age and gender, as well as specific data on HIV/AIDS. In particular, some DHS include an HIV serological survey, for which participants are asked to consent to be tested for HIV (anonymously in most of the surveys included here). Household wealth was assessed using the DHS wealth index, a composite measure of living standards that is based on the household’s assets (e.g. refrigerators) and characteristics (e.g., sanitation facilities) [13].

All DHS were conducted by national central statistics agencies or research institutes. The institutions that approved, implemented, or provided funding for the surveys were responsible for ethical clearance, which guaranteed informed consent and confidentiality of participant’s information.

### HIV indicators

We defined and analysed a set of eight HIV indicators aimed at capturing HIV knowledge, attitudes, behaviours and access to and uptake of HIV prevention services. These indicators were chosen and designed in a pragmatic way based on the information collected in the DHS across all countries selected for this analysis. These HIV indicators were consistently coded as binary variables with the level 1 (the level 0, respectively) reflecting a favourable (unfavourable, respectively) condition regarding HIV. These HIV indicators were:

i. *HIV-related knowledge*: based on the responses to a set of seven questions related to HIV transmission and prevention, we defined a categorical indicator reflecting good HIV-related knowledge (coded as 1 for those answering all questions correctly, 0 otherwise).
ii. *Positive attitudes towards PLHIV*: based on a set of two questions related to stigma towards PLHIV, we defined a categorical indicator reflecting positive attitudes towards PLHIV (coded as 1 for those answering positively to both questions, 0 otherwise).
iii. *No multipartnership (no or one sexual partner) in the past year*: based on the number of sexual partners reported in the last 12 months (self-reported, coded as 1 for those reporting no or one sexual partner, 0 otherwise)
iv. *Condom use at last sexual intercourse* (self-reported, coded as 1 for those reporting having used a condom during their last intercourse, 0 otherwise).
v. *VMMC* (among men only): based on the self-reported circumcision status and the qualification of the person who performed the circumcision (health worker/professional). We only analysed VMMC in 7 countries among of 14 that have been prioritized for the implementation of VMMC as HIV-prevention intervention in 2007 (mainly in Eastern and Southern Africa) [14].
vi. *Participation in Prevention of Mother to Child Transmission (*PMTCT, among women only): based on self-reported HIV testing during antenatal care among women who gave birth in the 12 months preceding the survey (coded as 1 for those reporting having been tested, 0 otherwise).
vii. *Recent HIV testing*: based on self-reported HIV testing within the past 12 months (coded as 1 for those reporting having been recently tested, 0 otherwise).
viii. *HIV seronegativity among youth*: based on the result of the HIV serological survey among 15-24 years old participants (coded as 1 in seronegative youths, 0 otherwise). trends in HIV prevalence among youth (aged 15-24 years) have been documented to be a surrogate reflecting trends in HIV incidence [15,16]. As the onset of sexual activity in this group is likely to be recent, prevalence reflects recent infection and therefore incidence. We thus used the socioeconomic gradient in HIV seroprevalence among the 15-24 years old as a proxy for the socioeconomic gradient in HIV incidence. To consistently consider a level 1 reflecting a favourable condition and therefore facilitate the interpretation and comparison of inequalities across indicators, the level 1 was chosen to code HIV seronegativity.

Further details on the construction of these HIV-related indicators are provided in Appendix 1

### Statistical analysis

For each country survey and each HIV indicator, we calculated percentages of positivity taking into account the survey design and sampling weights. We then assessed within-country socioeconomic inequalities related to each indicator based on participants’ relative rank in the cumulative distribution of the wealth index. Inequalities were measured on both relative and absolute scales using the relative index of inequality (RII) and the slope index of inequality (SII), respectively [17]. The RII expresses the ratio of the predicted outcomes between the richest and the poorest people in the wealth distribution. The SII represents the absolute difference in the predicted proportions of these two extremes. For instance, a RII value of 2 indicates that individuals at the upper extreme of the wealth distribution (the richest) are twice more likely to report the outcome than individuals at the lower extreme (the poorest). A SII value of 0.2 means that the proportion of the outcome is higher by 0.2 (or higher by 20 percentage-points) in the upper extreme of the wealth distribution (the richest) versus the lower extreme (the poorest). Reporting inequalities on both scales is recommended, because conclusions can be skewed when only one or the other is used [18]. Furthermore, the choice of a relative scale over an absolute scale or vice versa carries an implicit normative judgment on what a fair and socially just distribution of health should be [19].

Both RII and SII were obtained by fitting a modified Poisson regression with a robust variance and a log link function to estimate the association between participants’ relative wealth rank and each indicator, and by using generalised estimating equations to account for the clustering of observations [20]. We used the Wilcoxon rank-sum test to compare indices of inequalities between West and Central Africa (WCA) versus East and Southern African (ESA) countries (except for *VMMC*, which was studied in ESA only). Lastly, we used random-effects meta-analysis to average inequality estimates across countries [21].

## Results

### Study population

DHS included in the present work were conducted between 2010 and 2018 (Figure 1), with participation rates ranging from 90% (Ethiopia 2016) to 100% (Rwanda 2014-15) (Table 1). Overall, these studies represented 358,591 participants, (238,055 women and 120,536 men).

**Table 1:** Distribution of HIV-related indicators for 18 Sub-Saharan African countries (Demographic and Health Surveys). PLHIV: People living with HIV; PMTCT : Prevention of mother-to-Child Transmission; VMMC: Voluntary Medical Male Circumcision; NA: Not Available. ^1^: as reported in the country’s DHS final report; ^2^: within the past year; ^3^: at last sexual intercourse; ^4^: among those aged 15-24 y. Countries are ordered west to east.

| West and Central Africa |  |  |  |  |  |  |  |  |  |  |
| --- | --- | --- | --- | --- | --- | --- | --- | --- | --- | --- |
|  | Senegal<br>(2017) | Sierra Leone<br>(2013) | Guinea<br>(2018) | Liberia<br>(2013) | Côte d'Ivoire<br>(2011-12) | Mali (2012-13) | Burkina Faso<br>(2010) | Niger<br>(2012) | Cameroon<br>(2018) | DR Congo<br>(2013-14) |
| Response rate (%)<sup>1</sup> | 92% | 96% | 98% | 96% | 92% | 94% | 97% | 91% | 97% | 98% |
| Sample size | 23 764 | 23 920 | 14 991 | 13 357 | 15 195 | 14 823 | 24 394 | 15 088 | 20 505 | 27 483 |
| HIV-related knowledge | 17.7% | 19.5% | 15.8% | 6.2% | 14.0% | 19.9% | 22.7% | 11.0% | 30.3% | 12.5% |
| Positive Attitude towards<br>PLHIV | 32.9% | 34.1% | 16.0% | 34.7% | 47.0% | 45.1% | 34.5% | 20.1% | 55.1% | 35.7% |
| No multipartnership<sup>2</sup> | 97.1% | 88.1% | 95.4% | 90.1% | 88.1% | 95.2% | 94.2% | 95.6% | 90.1% | 91.1% |
| Condom use<sup>3</sup> | 6.5% | 5.0% | 8.3% | 12.8% | 18.5% | 3.6% | 12.7% | 1.1% | 29.7% | 9.2% |
| Participation to PMTCT<br>(Females) | 67.0% | 72.2% | 30.7% | 76.4% | 50.0% | 23.2% | 40.3% | 33.5% | 89.4% | 25.0% |
| VMMC (Males) | - | - | - | - | - | - | - | - | - | - |
| Recent HIV-Testing<sup>2</sup> | 18.6 | 14.7% | 8.4% | 19.1% | 13.5% | 6.8% | 10.3% | 6.9% | 57.7% | 8.6% |
| HIV-seronegativity among<br>youth<sup>4</sup> | 99.8% | 98.9% | 99.1% | 98.9% | 98.7% | 99.2% | 99.7% | 100% | 98.7% | 99.3% |
| East and Southern Africa |  |  |  |  |  |  |  |  |  |  |
|  | Zambia<br>(2018) | Lesotho<br>(2014) | Zimbabwe<br>(2015) | Rwanda<br>(2014-15) | Malawi (2015-16) | Tanzania<br>(2011-12) | Kenya<br>(2014) | Ethiopia<br>(2016) |  |  |
| Response rate (%)<sup>1</sup> | 94% | 95% | 94% | 100% | 96% | 97% | 92% | 90% |  |  |
| Sample size | 25 815 | 9 552 | 18 351 | 19 714 | 32 040 | 19 319 | 11 909 | 28 371 |  |  |
| HIV-related knowledge | 29.6% | 23.8% | 40.2% | 43.4% | 34.7% | 28.3% | 27.2% | 22.2% |  |  |
| Positive Attitude towards<br>PLHIV | 72.8% | 80.5% | 77.9% | 84.3% | 81.6% | 61.5% | 75.2% | 39.7% |  |  |
| No multipartnership<sup>2</sup> | 92.1% | 87.6% | 92.9% | 98.0% | 96.1% | 88.9% | 93.4% | 98.2% |  |  |
| Condom use<sup>3</sup> | 18.8% | 51.3% | 23.4% | 12.4% | 14.6% | 19.3% | 21.6% | 3.0% |  |  |
| Participation to PMTCT<br>(Females) | 97.3% | 98.4% | 97.9% | 99.4% | 97.2% | 80.5% | 98.0% | 57.2% |  |  |
| VMMC (Males) | 40.4% | NA | 11.3% | 85.4% | 9.0% | 40.7% | 60.0% | 15.8% |  |  |
| Recent HIV-Testing<sup>2</sup> | 59.8% | 52.6% | 43.6% | 38.8% | 43.9% | 30.4% | 64.9% | 20.6% |  |  |
| HIV-seronegativity among<br>youth<sup>4</sup> | 96.1% | 90.3% | 95.0% | 99.0% | 96.9% | 98.0% | NA | 99.8% |  |  |

**Figure 1:**
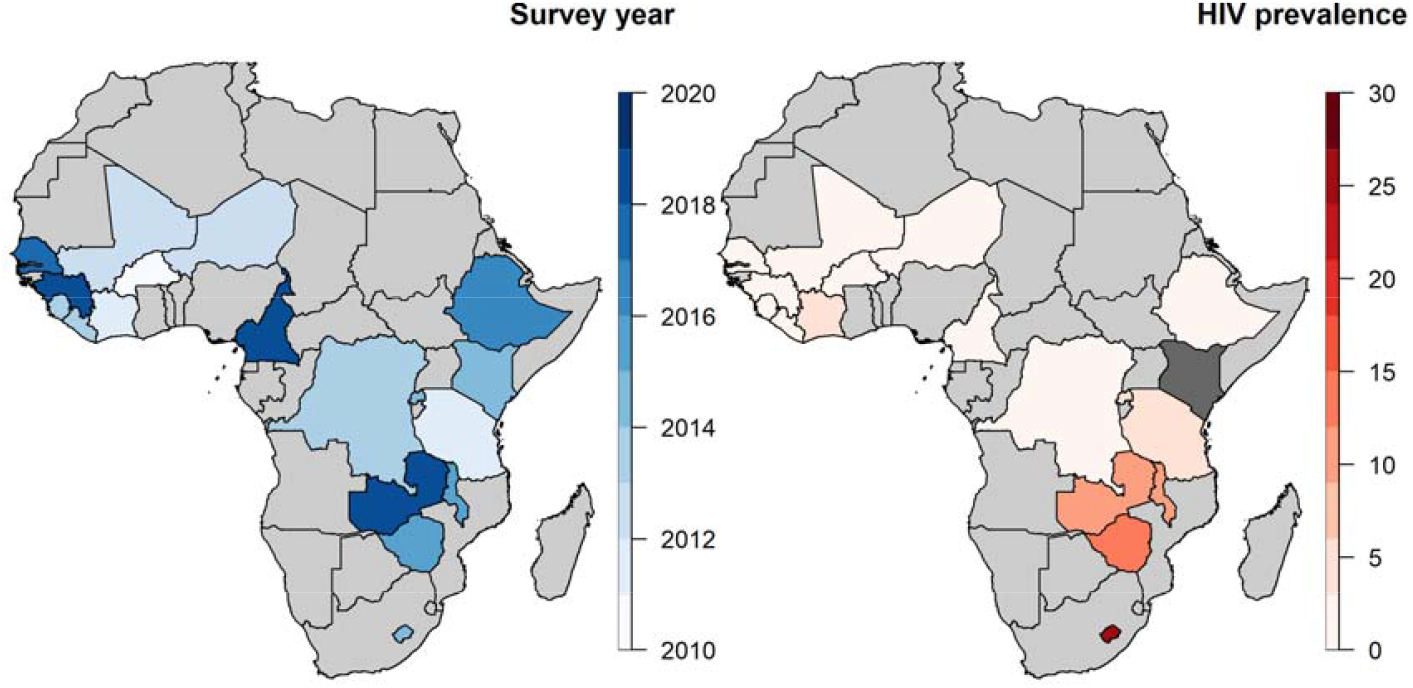
Demographic and Health Survey (DHS) year (left) and adult HIV prevalence (right) of the 18 Sub-Saharan African countries. (NB: For Kenya, HIV prevalence is unavailable in its last DHS 2014).

### HIV indicators

Country-specific percentages of positivity for each HIV indicator are presented in Table 1. In ESA countries, good HIV-related knowledge ranged from 22% (Ethiopia) to 43 % (Rwanda) while it was ≤20% in 8 out of 10 WCA countries (with the exception of Cameroon, 30% and Burkina Faso, 23%). Similarly, positive attitudes toward PLHIV were >60% in 7 out of 8 ESA countries (with the exception of Ethiopia, 40%) while the highest level in WCA was 55% (Cameroon). Reporting no multipartnership in the past 12 months was consistently high across countries, ranging from 88% (Sierra Leone, Côte d’Ivoire and Lesotho) to 98% (Rwanda and Ethiopia). Reported condom use at last sexual intercourse was highest in Lesotho (51%). In other countries reported condom use was lower, ranging from 1% (Niger) to 23% (Zimbabwe). Overall, higher uptake of HIV testing in the past 12 months was observed in ESA, with uptake >20% in all countries, as compared to in WCA, where uptake was <20% in 9 out of 10 countries. Similarly, ESA countries reported levels of participation to PMTCT >95% in 6 out of 8 countries (with the exception of Ethiopia, 57% and Tanzania, 81%), while it was <90% in all WCA countries, with large heterogeneity (from 23% in Mali to 89% in Cameroon). As mentioned earlier, VMMC was reported in ESA countries only, with levels ranging from 9% (Malawi) to 85% (Rwanda).

### Socioeconomic inequalities in the HIV indicators

Measures of relative and absolute inequalities pooled across countries are presented in Table 2. Significant levels of relative and absolute inequalities in favour of the richest were observed for HIV-related knowledge, positive attitudes toward PLHIV, condom use at last sex, participation to PMTCT, VMMC and recent HIV testing. In contrast, we observed inequalities in the other direction in reporting no multipartnership and in HIV-seronegativity among youth (that is, the poorest reported lower levels of multipartnership and exhibited lower levels of HIV seropositivity), although these inequalities were borderline significant and of low magnitude as compared to the other indicators. The ranking of the indicators by level of inequalities varied between the relative and the absolute scales. On the relative scale, the highest level was observed for condom use, with the richest being approximatively 5 times more likely (RII = 5.02; 95 Confidence interval, CI, 2.79-9.05) to use condom than the poorest people. In the absolute scales, the largest gap between the richest and the poorest people was observed in positive attitudes toward PLHIV, with a difference of 32 percentage-points (SII=0.32; 95% CI 0.26-0.39).

**Table 2:** Pooled relative and absolute wealth-related inequalities in various HIV-related indicators in sub-Saharan Africa, per region and overall. RII: relative index of inequality; SII : slope index of inequality; PLHIV: People living with HIV; PMTCT : Prevention of Mother-to-Child Transmission; VMMC: Voluntary Medical Male Circumcision. ^1^: rank-sum test p-value; ^2^: within the past year; ^3^: at last sexual intercourse; ^4^: among those aged 15-24 y.

|  | West and Central Africa |  | East and Southern Africa |  |  | Overall (all countries) |  |
| --- | --- | --- | --- | --- | --- | --- | --- |
|  | Pooled RII [95% CI] | I2 | Pooled RII [95% CI] | I2 | P-value<sup>1</sup> | Pooled RII [95% CI] | I2 |
| HIV-related knowledge | 3.13 [2.26;4.34] | 52% | 1.89 [1.33;2.69] | 70% | p = 0.0145 | 2.45 [1.91;3.14] | 72% |
| Positive Attitude towards PLHIV | 2.49 [1.74;3.56] | 78% | 1.60 [1.19;2.15] | 76% | p = 0.016 | 1.99 [1.57;2.53] | 81% |
| No multipartnership<sup>2</sup> | 0.99 [0.97;1.00] | 0% | 0.99 [0.98;1.01] | 0% | p = 0.32 | 0.99 [0.98;1.00] | 0% |
| Condom use<sup>3</sup> | 8.08 [3.87;16.87] | 77% | 2.80 [1.13;6.95] | 88% | p = 0.007 | 5.02 [2.79;9.05] | 88% |
| Participation to PMTCT (Females) | 2.87 [1.60;5.15] | 82% | 1.09 [0.90;1.33] | 51% | p = 0.002 | 1.70 [1.20;2.43] | 85% |
| VMMC (Males) | - | - | 2.08 [1.38;3.15] | 59% | - | - | - |
| Recent HIV-Testing<sup>2</sup> | 4.47 [2.13;9.40] | 90% | 1.24 [0.85;1.81] | 76% | p = 0.003 | 2.43 [1.47;4.04] | 92% |
| HIV-seronegativity among youth<sup>4</sup> | 0.99 [0.98;0.99] | 0% | 0.98 [0.95;1.02] | 0% | p = 0.45 | 0.99 [0.98;1.00] | 0% |
|  | West and Central Africa |  | East and Southern Africa |  |  | Overall (all countries) |  |
|  | Pooled SII [95% CI] | I2 | Pooled SII [95% CI] | I2 | P-value<sup>1</sup> | Pooled SII [95% CI] | I2 |
| HIV-related knowledge | 0.18 [0.13;0.23] | 98% | 0.18 [0.11;0.25] | 98% | p = 0.89 | 0.18 [0.14;0.22] | 98% |
| Positive Attitude towards PLHIV | 0.33 [0.23;0.43] | 99% | 0.31 [0.19;0.44] | 100% | p = 0.76 | 0.32 [0.26;0.39] | 99% |
| No multipartnership<sup>2</sup> | -0.02 [-0.04;-0.00] | 98% | -0.01 [-0.02;0.01] | 96% | p = 0.32 | -0.01 [-0.03;-0.00] | 97% |
| Condom use<sup>3</sup> | 0.17 [0.11;0.23] | 98% | 0.15 [0.10;0.20] | 96% | p = 0.69 | 0.16 [0.12;0.20] | 98% |
| Participation to PMTCT (Females) | 0.43 [0.31;0.55] | 97% | 0.12 [-0.07;0.32] | 99% | p = 0.009 | 0.29 [0.17;0.42] | 99% |
| VMMC (Males) | - | - | 0.20 [0.11;0.29] | 97% | - | - | - |
| Recent HIV-Testing<sup>2</sup> | 0.16 [0.11;0.21] | 97% | 0.09 [-0.01;0.19] | 99% | p = 0.06 | 0.13 [0.08;0.18] | 99% |
| HIV-seronegativity among youth<sup>4</sup> | -0.01 [-0.02;-0.01] | 92% | -0.01 [-0.05;0.03] | 98% | p = 1.00 | -0.01 [-0.03;0.00] | 97% |

On the relative scale, levels of inequalities were significantly higher in WCA as compared to ESA countries for the following indicators: HIV-related knowledge, positive attitude towards PLHIV, condom use, participation to PMTCT and recent HIV testing (all rank-sum test p-values <0.05). Absolute levels of inequalities were also higher in WCA versus ESA countries for participation to PMTCT (0.43 versus 0.12, respectively, p=0.009) and for recent HIV testing, although this was borderline significant (0.16 versus 0.09, respectively, p=0.06).

A more detailed comparison of the levels of inequalities across indicators and across countries is displayed in Figure 2. In WCA countries, the countries exhibiting the higher levels of inequalities were generally not the same when considering the relative or the absolute scale. In contrast, in ESA countries, Ethiopia tended to exhibit the highest levels of inequalities on both the relative and absolute scales. We also observed significant inequalities in favour of the poor in HIV-seronegativity among youth in some countries, especially on the absolute scale (for example in Sierra Leone, Zambia, Rwanda), although their magnitudes were lower than for other indicators.

**Figure 2:**
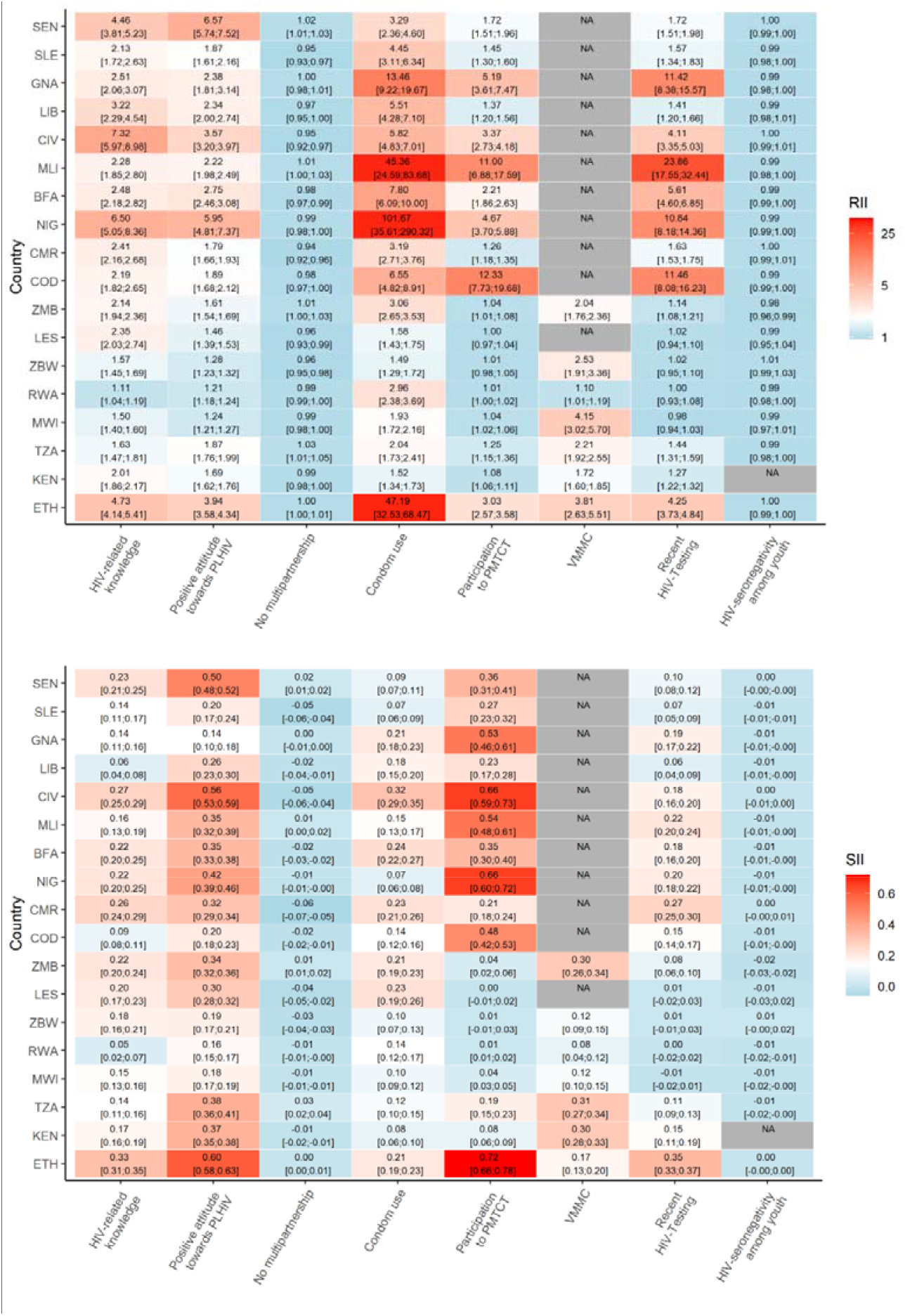
Relative (top) and absolute (bottom) wealth-related inequalities in various HIV-related indicators across 18 sub-Saharan African countries. Countries are ordered wet to east. RII: relative index of inequality; SII : slope index of inequality; PLHIV: People living with HIV; PMTCT : Prevention of Mother-to-Child Transmission; VMMC: Voluntary Medical Male Circumcision.

Table 3 presents the correlations between RII (log-transformed) and SII values across HIV-related indicators. We observed significant positive correlations between relative- and absolute inequalities metrics for most of the indicators (Pearson correlation coefficients >0.6 in 6 out of 8 indicators), apart from condom use and VMMC presenting no evidence of correlation.

**Table 3:** Correlation between country-level relative and absolute wealth-related inequalities, per HIV indicator. RII: relative index of inequality; SII: slope index of inequality; PLHIV: People living with HIV; PMTCT : Prevention of Mother-to-Child Transmission; VMMC: Voluntary Medical Male Circumcision. 1: log-transformed; 2: within the past year; 3: at last sexual intercourse; 4: among those aged 15-24 y.

|  | Correlation between<br>RII<sup>1</sup> and SII values, |  |
| --- | --- | --- |
|  | Pearson r | p-value |
| HIV-related knowledge | 0.607 | 0.0075 |
| Positive Attitude towards PLHIV | 0.716 | <0.001 |
| No multipartnership<sup>2</sup> | 0.992 | <0.001 |
| Condom use<sup>3</sup> | 0.064 | 0.7996 |
| Participation to PMTCT (Females) | 0.809 | <0.001 |
| VMMC (Males) | -0.098 | 0.8347 |
| Recent HIV-Testing<sup>2</sup> | 0.641 | 0.0041 |
| HIV-seronegativity among youth<sup>4</sup> | 1.000 | <0.001 |

## DISCUSSION

In this study, relying on cross-sectional population-based surveys, we provide an extensive assessment of socio-economic inequalities in various HIV indicators capturing knowledge, attitudes, behaviours and access to and uptake of prevention services in a large set of sub-Saharan African countries. We document important levels of wealth-related inequalities, both on the relative and absolute scales, in HIV-related knowledge, positive attitudes toward PLHIV, condom use, participation to PMTCT, uptake of VMMC and recent HIV testing. The magnitude of these inequalities varies across countries and indicators. However, inequalities tend to be more marked in WCA than in ESA countries. In contrast, we document low overall levels of inequalities in multi-partnership and HIV seropositivity among youth, a surrogate of HIV incidence.

### The arithmetic behind inequality measures

The results we provide here allow two types of comparisons: cross-country comparisons of inequalities given a specific HIV indicator, and within-country comparisons of inequalities across HIV indicators. However, first bearing in mind some arithmetic considerations is important when comparing RII and SII across indicators and countries [22,23]. Indeed, for any indicator, the overall level in the population should be considered when interpreting relative and absolute inequalities measures. Relative inequalities tend to be larger at low overall levels of the considered indicator, while absolute inequalities tend to be larger at intermediate levels. This arithmetic explains, for instance, why condom use in Niger (overall self-reported level: 1.1%) acknowledges dramatic relative inequalities (RII >100) but relatively limited absolute inequalities (SII = 0.07). This illustrates the importance of reporting both relative and absolute measures when reporting inequalities, as conclusions would be quite different if based on only one or the other indicators in this single example. However, our analysis of correlation showed that the levels of inequalities tended to correlate across both scales for most of the indicators we used.

### Cross-country comparisons of inequalities

Heterogeneity across countries regarding the inequalities reported here may be driven by a potentially large spectrum of socioeconomic, epidemiological or healthcare-related factors. Nevertheless, we observed that geography explained some of this heterogeneity: higher levels of wealth-related inequalities were observed in countries in WCA compared to those in ESA, especially on the relative scale. Several factors may explain higher levels of inequalities in WCA. This may be related to the historical prominence of the private healthcare sector in this region, which generally implies user fees that may act as a barrier to access to care for the poorest [24]. Higher levels of inequalities are also possibly linked to the smaller overall HIV epidemics in WCA compared to ESA. Indeed, ESA countries had higher HIV prevalence than WCA countries in our sample, except for Ethiopia, which precisely had the highest levels of wealth-related inequalities within ESA countries. Previous results suggested that HIV-related inequalities correlated with epidemiological rather than macro-economic factors [25]. In addition, the country-level HIV prevalence has been reported to correlate with levels of HIV donor and national funding for HIV prevention services [26]. Moreover, donor funding tended to prioritise prevention programs targeting key populations in WCA, as new HIV infections are often predominantly in these populations in the region, whilst in ESA HIV prevention efforts are broader encompassing both key populations and the general populations [27]. While key populations deserve a priority focus for HIV prevention in WCA, additional efforts should be considered to overcome the socioeconomic disparities in knowledge and access to HIV testing and prevention services more broadly.

### Comparisons of inequalities across HIV indicators

Overall, we did not observe large socioeconomic disparities in reporting having multiple sexual partners, consistently with recent results [28]. Large absolute and relative inequalities, overall, remain regarding lack of knowledge and stigmatizing attitudes toward PLHIV, and this may undermine HIV prevention, care and treatment [29]. Concerning inequalities are observed in condom use, PMTCT, VMMC and HIV testing, especially in WCA countries. This is of particular concern because these are interventions that can prevent new infections, directly for condom use and VMMC, or when linked to care and treatment for PMTCT and HIV testing. Large inequalities in condom use may reflect difficulties to access condoms free of charge. However, PMTCT, VMMC and HIV testing are also interventions that are usually provided at no direct cost for the individual, which underlines that providing these services without cost to clients is not the only factor to ensure equitable access to prevention interventions. Lessons should be drawn from the experiences of PMTCT or HIV testing programs in ESA countries that are currently offered to all at no cost, without producing measurable health inequalities. For instance, Rwanda has successfully integrated HIV services within the existing healthy system and assured delivery of services in remote areas. These strategy have strengthened the country response to HIV and have most probably contributed to limiting health inequalities [30].

Contrary to what one could expect considering the inequalities disfavouring the poorest in terms of access to HIV prevention service that we report here, we did not find any evidence of socioeconomic inequalities regarding HIV prevalence among the youth, an indicator we used as a proxy for HIV incidence. This apparently inconsistent result may however be linked to the complex and changing social epidemiology of HIV. Many early studies reported that higher SEP tended to be associated with a higher prevalence of HIV infection, making this distribution unusual as compared to many health outcomes [31,32]. The absence of overall socioeconomic gradient we report here may represent a transient state were inequalities are inversing, from a higher risk for the wealthiest to a higher risk for the poorest [33,34]. Indeed, a recent study relying on a >20-years follow-up among a population-based open cohort in rural Uganda documented a widening socioeconomic gradient over time, with a higher risk of incident HIV infection among the poorest [35]. An effort to more systematically monitor socio-economic inequalities in HIV incidence is thus of high importance to detect possibly changing SEP gradients. It is anticipated that the newer-generation surveys, such as the population-based HIV impact assessment surveys (https://phia.icap.columbia.edu/) which integrate HIV incidence assays, may provide useful data for this monitoring.

### Main strengths and limitations of the study

This study has several limitations and strengths. The between-country comparison may be affected by the period of data collection for each national survey (from 2010 to 2018). Large HIV programs have been implemented over this study period and may have reduced inequalities over time [9]. Our analysis was limited to indicators pragmatically built on data collected as part of the DHS. As such, it matches only partially the recently proposed unifying framework of the HIV prevention cascade [36]. For instance, an important aspect absent from the DHS data, and therefore from our analysis, relates to knowledge and use of antiretroviral treatment for individual and collective health benefits. For the same reason, our analysis ignores important prevention interventions, such as pre-exposure prophylaxis (PrEP), or interventions targeted to key populations. However, to our knowledge, this study constitutes the first effort to quantify both relative and absolute socioeconomic inequalities on a large set of HIV-related indicators collected from large, representative surveys conducted in numerous SSA countries.

## Conclusions

In conclusion, this study provides a comprehensive monitoring of socioeconomic inequalities in HIV knowledge, attitudes, behaviours and prevention in sub-Saharan Africa. In other fields such as child health, such monitoring has provided important insight in the way policies may be tailored to the patterns of inequalities in order to best address them [37]. We hope that this study will help in the strategical articulation of HIV prevention approaches that is required for fulfilling the focus on reducing inequalities that the renewed 2021-2026 Global AIDS Strategy adopted [38].

## Data Availability

Data referred to in the manuscript are available at www.dhsprogram.com

https://www.dhsprogram.com/

## Authors’ contributions

MH, PAA-T and KJ conceived and planned the study with input from LT. MH and PAA-T collated and processed DHS data. MH and KJ conducted the analysis and produced output figures and tables with input from PAA-T and LT. All authors contributed to the interpretation of the results. MH and KJ wrote the first draft of the report and all authors contributed to subsequent revisions.

